# *Salmonella* Typhi Haplotype 58 (H58) Biofilm Formation and Genetic Variation in Typhoid Fever Patients with Gallstones in an Endemic Setting in Kenya

**DOI:** 10.1101/2024.06.03.24308409

**Authors:** Peter Muturi, Peter Wachira, Maina Wagacha, Cecilia Mbae, Susan Kavai, Michael Mugo, Musa Muhammed, Juan F. González, Samuel Kariuki, John S. Gunn

## Abstract

The causative agent of typhoid fever, *Salmonella enterica* serovar Typhi, is a human restricted pathogen. Human carriers, 90% of whom have gallstones in their gallbladder, continue to shed the pathogen after treatment. The genetic mechanisms involved in establishing the carrier state are poorly understood, but *S*. Typhi is thought to undergo specific genetic changes within the gallbladder as an adaptive mechanism. In the current study, we aimed to identify biofilm forming ability and the genetic differences in longitudinal clinical *S*. Typhi isolates from asymptomatic carriers with gallstones in Nairobi, Kenya. Whole genome sequences were analyzed from 22 *S*. Typhi isolates, 20 from stool and 2 from blood samples, all genotype 4.3.1 (H58). Nineteen strains were from four patients also diagnosed with gallstones, of whom, three had typhoid symptoms and continued to shed *S*. Typhi after treatment. All isolates had point mutations in the quinolone resistance determining region (QRDR) and only sub-lineage 4.3.1.2EA3 encoded multidrug resistance genes. There was no variation in antimicrobial resistance patterns among strains from the same patient/household. Non-multidrug resistant (MDR), isolates formed significantly stronger biofilms *in vitro* than the MDR isolates*, p<0.001*. A point mutation within the *treB* gene (*treB* A383T) was observed in strains isolated after clinical resolution from patients living in 75% of the households. Missense mutations in Vi capsular polysaccharide genes, *tviE* P263S was also observed in 18% of the isolates. This study provides insights into the role of typhoid carriage, biofilm formation, AMR genes and genetic variations in *S.* Typhi from asymptomatic carriers.

**Importance:** Although typhoid fever has largely been eliminated in high income countries, it remains a major global public health concern especially among low- and middle-income countries. The bacteria responsible for this infectious disease, *Salmonella* Typhi, has limited ability to replicate outside the human host and human carriers serve as a reservoir of infection. Typhoid is a common infection in parts of sub-Saharan Africa and Asia, and is endemic in our study setting. Our research findings on differences in *S.* Typhi strains causing typhoid fever and carriage will influence public health approaches aimed at reducing carriage and transmission of *S*. Typhi.

## Introduction

Typhoid fever (typhoid), a life-threatening systemic infection caused predominantly by *Salmonella enterica* subspecies *enterica* serotype Typhi (*S*. Typhi), remains a common infection and a public health concern in resource poor-settings in parts of sub-Saharan Africa and Asia (1–3). An estimated 9 million new typhoid fever cases occur each year, of which 2-3% results in death even with adequate antibiotics therapy (4,5). Typical symptoms manifest between 1 and 3 weeks post-infection and encompass elevated prolonged fever, headache, malaise, abdominal pain, diarrhea, constipation, hypersplenism and rose-colored spots on the chest (6). *S*. Typhi is transmitted via the fecal-oral route in settings with poor standards of sanitation, low levels of hygiene and inadequate water supply (7,8). Besides inadequate resources, typhoid endemic settings lack a quality public health infrastructure (9).

Upon ingestion of contaminated food or water, *S.* Typhi bacteria that survive the hostile gastric acid rich environment in the stomach, are able to replicate in the new host (9,10). The typhoid bacilli can invade the intestinal mucosa, typically through microfold (M) cells, and establish an initially clinically undetectable infection involving significant systemic dissemination and a transient primary bacteremia (9). *S*. Typhi also reach the gallbladder hematogenously during primary bacteremia or shortly thereafter through infected hepatic bile entering the gallbladder (11,12). *S*. Typhi bacteria can survive, replicate and evade immune surveillance intracellularly within a modified phagosome known as *Salmonella*-containing vacuole (SCV) (13,14).

Although the majority of patients recover from typhoid fever after an appropriate treatment, some individuals become asymptomatic carriers and shed the infectious typhoid bacilli intermittently in their faeces for an ill-defined period of time after apparent clinical resolution. Since the early 20^th^ century, asymptomatic carriage has been demonstrated to be a source of transmission of typhoid fever, including in the famous case of Mary Mallon (15). Generally, ∼2–5% of acute typhoid cases fail to clear the infection fully within one year and develop asymptomatic chronic carriage (16–18). Approximately 90% of typhoid chronic carriers have gallstones in their gallbladder (19,20). Persistent colonization of the gallbladder by *S*. Typhi is facilitated by formation of biofilms on the surface of cholesterol gallstones (19,21). Biofilms are organized three-dimensional multicellular communities encased in self-produced extracellular polymeric substances (EPS) comprised of polysaccharides, extracellular DNA [eDNA], proteins and lipids (22). Biofilms account for 80% of chronic infections in humans, leading to increased rates of hospitalization, high health care costs, and increased mortality and morbidity rates (23). Bacteria in a biofilm are protected from certain environmental stresses, such as osmotic shifts, oxidative stress, metal toxicity, dehydration, radiation, host immunity, antimicrobial agents, and disinfectants (24). Chronic *S*. Typhi colonization usually cannot be resolved with antibiotics; gallbladder resection is the only option, although not always effective (21). Biofilm formation leads to continuous shedding and reattachment of planktonic cells, followed by bacteria diffusion in urine and feaces (19,25). Since *S*. Typhi is a human-restricted pathogen, gallbladder colonization and fecal shedding form a central dogma for further transmission and persistence of typhoid fever.

In the gallbladder, *S*. Typhi is exposed to bile, a complex digestive secretion comprised of bile acids, bilirubin, phospholipids and cholesterol, that exhibit strong antimicrobial properties (26,27). The molecular mechanisms involved in establishing the carrier state are poorly understood; however *S*. Typhi is thought to undergo genetic changes within the gallbladder as an adaptive mechanism (21,28,29).

Although it is widely accepted that *S*. Typhi carriers contribute to typhoid transmission in endemic settings, little progress has been made in understanding typhoid carrier state. The current study, we aimed at identifying the genetic differences in longitudinal clinical *S*. Typhi isolates from carriers, in a typhoid endemic setting in Nairobi, Kenya.

## RESULTS

### Genotype Identification and Clustering Tree

Since the *S*. Typhi population is highly structured, with dozens of subclades being associated with specific geographical regions and antimicrobial resistance patterns, the genotypes causing asymptomatic carriage in the current study settings were identified. All 22 bacteria isolates were genotype 4.3.1 (*S.* Typhi Haplotype 58 [H58]). Within this genotype, 11 of the isolates (from 2/4 of the households) fell under the lineage 4.3.1.1 (from households A and B) while the remaining 11 isolates (from households C and D) belong to lineage 4.3.1.2 (Figure 1). Lineage 4.3.1.2 strains were further grouped into two sub-lineages, 4.3.1.2EA2 (4 isolates from the index case in household C) and 4.3.1.2EA3 (7 isolates from an index case in household D). Each patient shed typhoid bacilli belonging to only one lineage/sub-lineage. From household A, isolates (i)-(v) were from and index case while isolate (vi) was from a household contact. Isolates (i) and (ii), household B, were from index case while (iii), (iv) and (v) were from an asymptomatic household contact living with the index case. All the *S*. Typhi strains were isolated from stool samples apart from two, isolate (i) from household B, and isolate (i) from household D, which were isolated from blood samples. The time of isolation/shedding of each isolate is shown in Table 1. Assembled genomes can be accessed in NCBI database, BioProject ID PRJNA1101423, GenBank accession numbers are shown in the data availability section.

**Figure 1.**
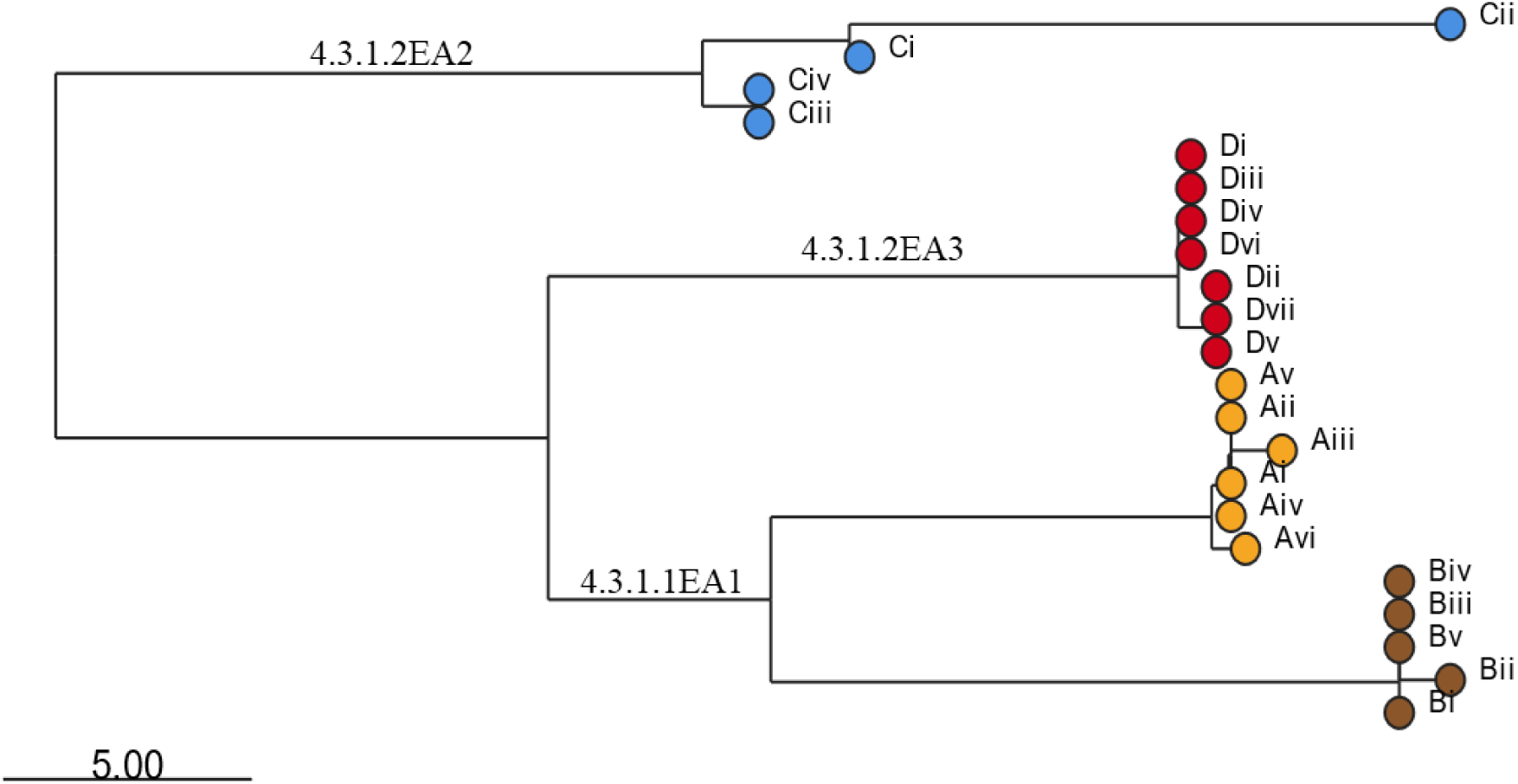
Clustering tree for *S.* Typhi strains isolated from the study participants living in the different households as generated using the pathogenwatch database and visualized using microreact. Roman numbers indicate specific isolates from the different households (A, B, C and D). Isolates from each household are shown by specific leaf nodes color. Branch tips are color-coded according to the household of isolation. The bar represents branch length scale bar. The tree can be visualized here https://microreact.org/project/s3biz8PE9LDnFayjco22MZ-s-typhi-kenya-2023.

**Table 1.**
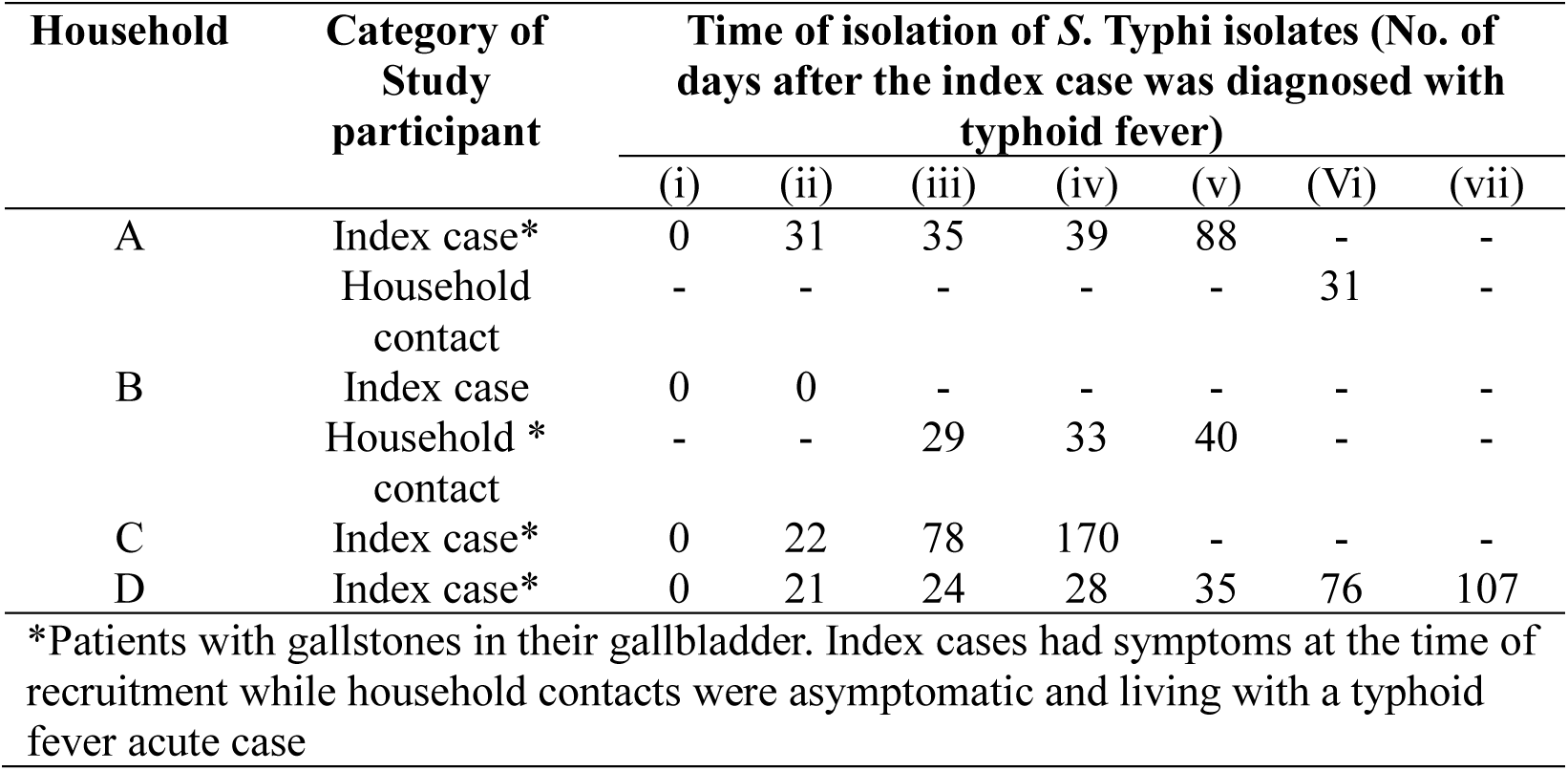
Time of isolation/shedding of *S.* Typhi.

### Antimicrobial Resistance Genes

Different antimicrobial resistance patterns were observed in the isolated *S*. Typhi strains. The seven isolates belonging to sub-lineage 4.3.1.2EA3 (from Household D) were multidrug resistant, all expressed the following acquired antimicrobial resistance genes; *sul1*, *dfrA7*, *catA1*, *aph*(*6*)*-Id*, *aph(3’’)-Ib*, *sul2* and *bla_TEM-1_*, and a point mutation in the Quinolone Resistance Determining Region (QRDR) of *gyrA* (*gyrA* S83Y). Phenotypic susceptibility data showed that these seven isolates were resistant to ampicillin, chloramphenicol, trimethoprim-sulfamethoxazole and nalidixic acid but non-susceptible to ciprofloxacin. The four sub-lineage 4.3.1.2EA2 isolates (*S*. Typhi strains from Household C) had a *gyrB* S464F mutation in the QRDR, and all were non-susceptible to nalidixic acid and ciprofloxacin according to phenotypic susceptibility results. The third group, lineage 4.3.1.1, had six strains (Household A isolates) with a *gyrB* S464F mutation, also demonstrating non-susceptibility to nalidixic acid and ciprofloxacin. The other five lineage 4.3.1.1 strains (household B isolates) had *gyrA* S83F mutations in the QRDR, and showed resistance to nalidixic acid and non-susceptibility to ciprofloxacin (Table 2).

**Table 2.**
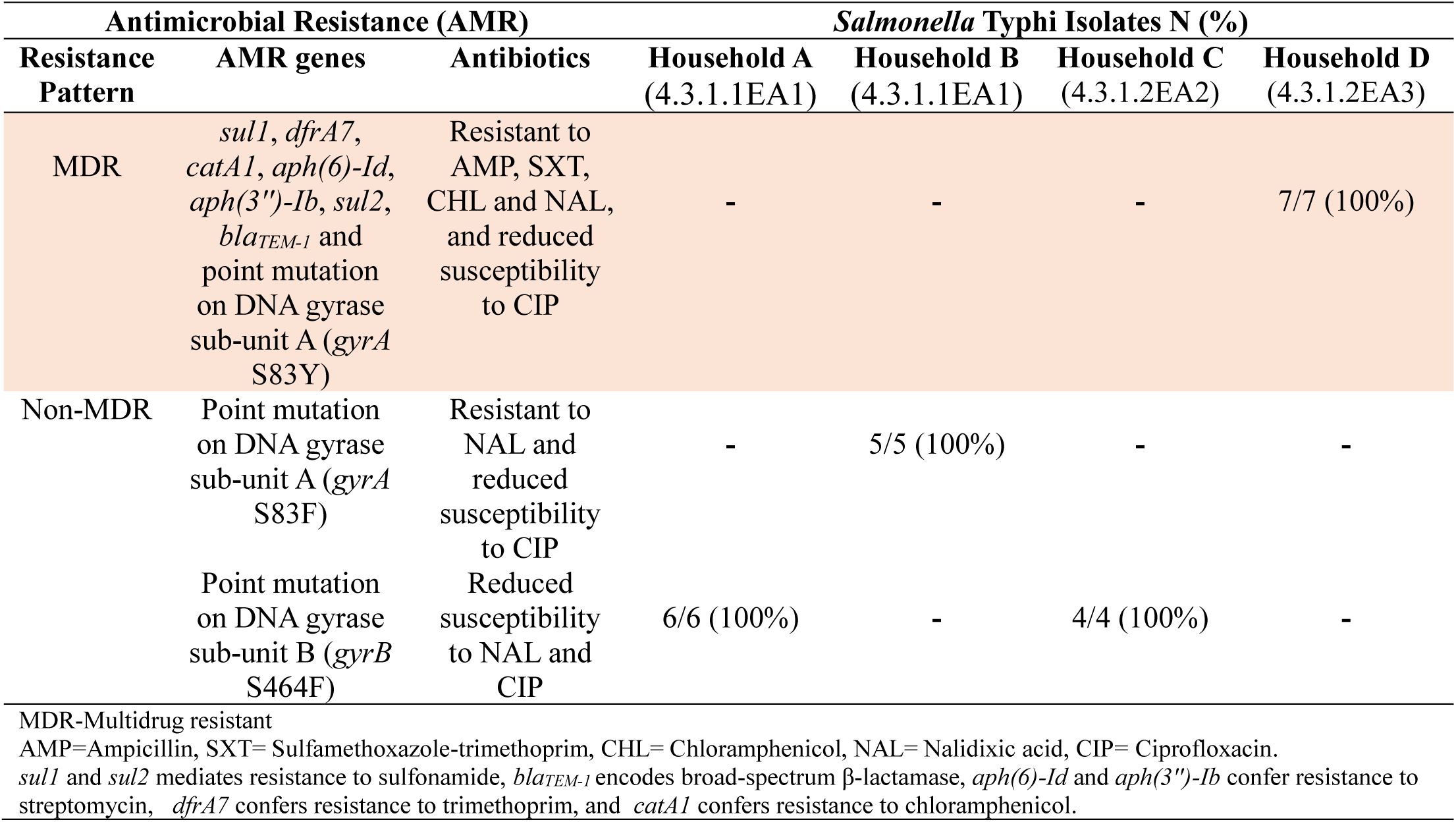
Antibiotic resistance profiles in isolated *S*. Typhi.

### Variant Calling

When comparing genome sequences among the isolates, missense mutations were observed in a gene that codes for Trehalose Phosphotransferase System (PTS), trehalose transporter subunit IIBC, *treB* gene (*treB* A383T), in 3/7 sub-lineage 4.3.1.2EA3 (Household D) isolates and in *tviE* gene (*tviE* P263S), which codes for the gene coding for biosynthesis of Vi polysaccharide biosynthesis protein TviE, in 4/7 isolates in the same group, while 2/7 had both mutations (*treB* A383T and *tviE* P263S), (Table 3). A nonsense mutation was observed in 1/7 of the sub-lineage 4.3.1.2EA3 isolates in the BCKKFA_12280 locus (E237*) that codes for integrase/transposase family protein. The first follow-up isolate (*S*. Typhi sub-lineage 4.3.1.2EA2) from the index case living in household C (C ii) had a total of 16 mutations, of which 7 were silent, 8 missense mutations and a deletion in *mutL* gene that codes for DNA mismatch repair endonuclease MutL (Table 3 and Supplementary files 2 and 3). The second and third follow-up isolates from the same patient (C iii and C iv) had two silent mutations, one in the *yccC* gene that codes for putative membrane protein YccC and *tehA* gene that codes for dicarboxylate transporter/tellurite-resistance protein TehA (*tehA* A124A and *yccC* R199R mutations respectively) and missense mutations in the *amiA* gene that codes for N-acetylmuramoyl-L-alanine amidase AmiA (*amiA* V145A mutation). An additional mutation was observed in the LEJNAJ_18700 locus (K124E) that encodes for 4-hydroxyphenylacetate permease. The second follow-up isolate from the patient in household C (C iii) had a third missense mutation in the LEJNAJ_10515 locus (D78G) that encodes phage baseplate assembly protein V. The follow-up lineage 4.3.1.1 strains, which were isolated from households A and B, also had a few mutations. While no mutation was detected in the first follow-up sample from the index case in household A, the second follow-up sample (A iii) had a single nucleotide polymorphism in the *crl* gene (*crl* L38P), *crl* codes for sigma factor-binding protein Crl. The third and fourth follow-up *S*. Typhi isolates from the same patient (A iv and A v) had a silent mutation in the *tnpA* gene (*tnpA* Y41Y) that codes for IS200/IS605 family transposase (Supplementary file 1). From the same household, an *S*. Typhi strain isolated from a household contact’s stool sample (A vi) had a missense mutation in the *treB* gene (*treB* A383T). *S*. Typhi bacteria isolated from the stool of the index case living in household B before treatment (isolate ii), had a missense mutation in the *waaK* gene (*waaK* P167L) that codes for lipopolysaccharide N-acetylglucosaminyltransferase, compared to the strain from blood sample collected on the same day (Supplementary file 2).

**Table 3.** Missense mutations in *S*. Typhi strains isolated from index cases after apparent clinical resolution and from asymptomatic household contacts.

| Gene/Locus | Seq Change | Household A<br>4.3.1.1EA1 |  |  |  | Household B<br>4.3.1.1EA1 |  |  |  | Household C<br>4.3.1.2EA2 |  |  | Household D<br>4.3.1.2EA3 |  |  |  |  |  |  |
| --- | --- | --- | --- | --- | --- | --- | --- | --- | --- | --- | --- | --- | --- | --- | --- | --- | --- | --- | --- |
|  |  | ii | iii | iv | v | vi | ii | iii | iv | v | ii | iii | iv | ii | iii | iv | v | vi | vii |
| *crL* | L38P |  | ✓ |  |  |  |  |  |  |  |  |  |  |  |  |  |  |  |  |
| *treB* | A383T |  |  |  |  | ✓ |  |  |  |  | ✓ |  |  | ✓ |  |  | ✓ |  | ✓ |
| *waaK* | P167L |  |  |  |  |  | ✓ |  |  |  |  |  |  |  |  |  |  |  |  |
| BCKKFA 12280 | E237* |  |  |  |  |  |  |  |  |  |  |  |  |  | ✓ |  |  |  |  |
| *tviE* | P263S |  |  |  |  |  |  |  |  |  |  |  |  |  |  | ✓ | ✓ | ✓ | ✓ |
| LEJNAJ 01195 | M224V |  |  |  |  |  |  |  |  |  | ✓ |  |  |  |  |  |  |  |  |
| *yhhY* | R73H |  |  |  |  |  |  |  |  |  | ✓ |  |  |  |  |  |  |  |  |
| LEJNAJ 13650 | R51W |  |  |  |  |  |  |  |  |  | ✓ |  |  |  |  |  |  |  |  |
| *yihT* | R270C |  |  |  |  |  |  |  |  |  | ✓ |  |  |  |  |  |  |  |  |
| *rfbP* | D196G |  |  |  |  |  |  |  |  |  | ✓ |  |  |  |  |  |  |  |  |
| *galR* | Y98H |  |  |  |  |  |  |  |  |  | ✓ |  |  |  |  |  |  |  |  |
| *dnaE* | E953G |  |  |  |  |  |  |  |  |  | ✓ |  |  |  |  |  |  |  |  |
| LEJNAJ 18700 | K124E |  |  |  |  |  |  |  |  |  |  | ✓ | ✓ |  |  |  |  |  |  |
| *amiA* | V145A |  |  |  |  |  |  |  |  |  |  | ✓ | ✓ |  |  |  |  |  |  |
| LEJNAJ 10515 | D78G |  |  |  |  |  |  |  |  |  |  | ✓ |  |  |  |  |  |  |  |

Roman numbers in red represent isolates from household contacts. ✓ indicates isolates with observed missense mutations.

### Plasmid Identification

*S*. Typhi genotype 4.3.1 lineages/sub-lineages identified in this study were found to contain plasmids. A total of 22 different plasmids were identified, 12/22 of these had sequences corresponding to known *Salmonella* species plasmids, 6/22 had sequences corresponding to *Escherichia coli* plasmids, 2/22 corresponding to *Klebsiella pneumoniae* plasmids, 1/22 corresponding to *Enterobacter kobei* and 1/22 an *Escherichia albertii* plasmid (Table 4). Out of the 22 plasmids, 8/22 were detected in all 22 *S*. Typhi strains. The eight plasmids include *Salmonella* Infantis strain CFSAN003307 plasmid pCFSAN003307, *Salmonella* Senftenberg strain NCTC10384 plasmid 4, *E. coli* strain 13TMH22 plasmid p13TMH22-1, *Salmonella* enterica subsp. enterica serovar 1,4,[5],12:i:-strain PNCS014880 plasmid p16-6773.2, *S.* Typhi strain ERL11909 plasmid 3, *Salmonella* Typhimurium strain AUSMDU00027951 isolate AUSMDU00027951 plasmid P02, *Salmonella* Weltevreden plasmid pSH17G0407 and *S.* Senftenberg strain NCTC10384 plasmid 3. Plasmids with sequences corresponding to *E. coli* strain 88COLEC plasmid p88COLEC-1 and *E. coli* strain 13P477T plasmid p13P477T-2 were only detected in lineage 4.3.1.2EA2 (Household C) isolates, in 4/4 and 2/4 of the strains respectively. The *E. albertii* strain 2014C-4356 plasmid unnamed6 was only found in 1/11 of lineage 4.3.1.1 isolates while *Salmonella* Uganda strain CFSAN006208 plasmid pCFSAN006208_1 was detected in 1/11 of lineage 4.3.1.1 isolates. Sequences corresponding to five known plasmids, *Salmonella* Enteritidis strain 81-1705 plasmid pSE81-1705-3, *S*. Typhi strain 311189_252186 plasmid pHCM1, *E. coli* NES6 plasmid pN-ES-6-1, *K. pneumoniae* strain RIVM_C018860 plasmid pRIVM_C018860_2 and *K.* pneumoniae strain 03108465-40B plasmid pKpQIL, were detected in 7/7 of sub-group 4.3.1.2EA3 strains but not in the other sub-groups.

**Table 4.**
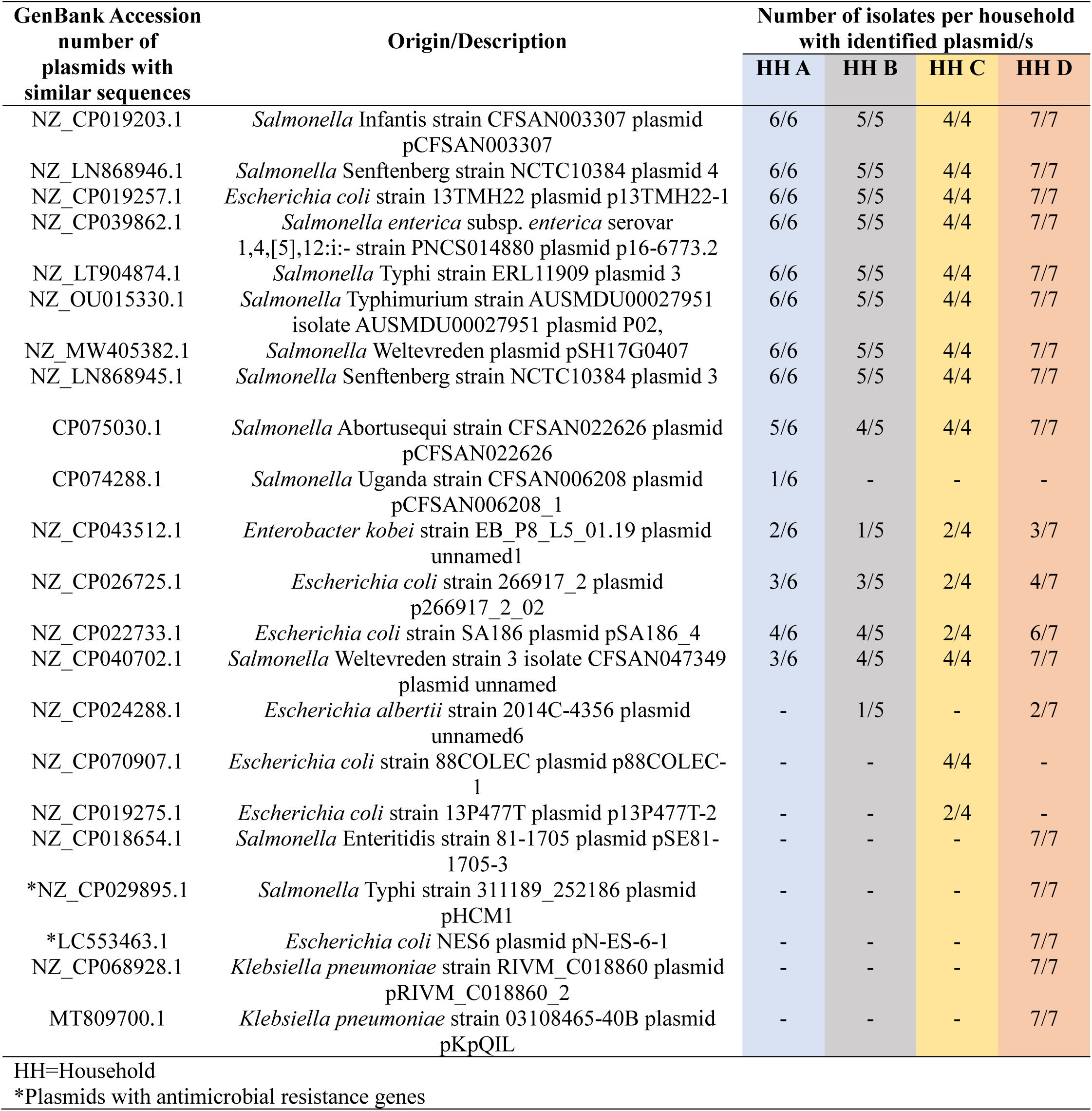
Bacterial plasmids detected in isolated *S.* Typhi strains.

| GenBank Accession<br>number of<br>plasmids with<br>similar sequences | Origin/Description | Number of isolates per household<br>with identified plasmid/s |  |  |  |
| --- | --- | --- | --- | --- | --- |
|  |  | HH A | HH B | HH C | HH D |
| NZ_CP019203.1 | *Salmonella* Infantis strain CFSAN003307 plasmid pCFSAN003307 | 6/6 | 5/5 | 4/4 | 7/7 |
| NZ_LN868946.1 | *Salmonella* Senftenberg strain NCTC10384 plasmid 4 | 6/6 | 5/5 | 4/4 | 7/7 |
| NZ_CP019257.1 | *Escherichia coli* strain 13TMH22 plasmid p13TMH22-1 | 6/6 | 5/5 | 4/4 | 7/7 |
| NZ_CP039862.1 | *Salmonella enterica* subsp. *enterica* serovar 1,4,[5],12:i:- strain PNCS014880 plasmid p16-6773.2 | 6/6 | 5/5 | 4/4 | 7/7 |
| NZ_LT904874.1 | *Salmonella* Typhi strain ERL11909 plasmid 3 | 6/6 | 5/5 | 4/4 | 7/7 |
| NZ_OU015330.1 | *Salmonella* Typhimurium strain AUSMDU00027951 isolate AUSMDU00027951 plasmid P02, | 6/6 | 5/5 | 4/4 | 7/7 |
| NZ_MW405382.1 | *Salmonella* Weltevreden plasmid pSH17G0407 | 6/6 | 5/5 | 4/4 | 7/7 |
| NZ_LN868945.1 | *Salmonella* Senftenberg strain NCTC10384 plasmid 3 | 6/6 | 5/5 | 4/4 | 7/7 |
| CP075030.1 | *Salmonella* Abortusequi strain CFSAN022626 plasmid pCFSAN022626 | 5/6 | 4/5 | 4/4 | 7/7 |
| CP074288.1 | *Salmonella* Uganda strain CFSAN006208 plasmid pCFSAN006208_1 | 1/6 | - | - | - |
| NZ_CP043512.1 | *Enterobacter kobei* strain EB_P8_L5_01.19 plasmid unnamed1 | 2/6 | 1/5 | 2/4 | 3/7 |
| NZ_CP026725.1 | *Escherichia coli* strain 266917_2 plasmid p266917_2_02 | 3/6 | 3/5 | 2/4 | 4/7 |
| NZ_CP022733.1 | *Escherichia coli* strain SA186 plasmid pSA186_4 | 4/6 | 4/5 | 2/4 | 6/7 |
| NZ_CP040702.1 | *Salmonella* Weltevreden strain 3 isolate CFSAN047349 plasmid unnamed | 3/6 | 4/5 | 4/4 | 7/7 |
| NZ_CP024288.1 | *Escherichia albertii* strain 2014C-4356 plasmid unnamed6 | - | 1/5 | - | 2/7 |
| NZ_CP070907.1 | *Escherichia coli* strain 88COLEC plasmid p88COLEC-1 | - | - | 4/4 | - |
| NZ_CP019275.1 | *Escherichia coli* strain 13P477T plasmid p13P477T-2 | - | - | 2/4 | - |
| NZ_CP018654.1 | *Salmonella* Enteritidis strain 81-1705 plasmid pSE81-1705-3 | - | - | - | 7/7 |
| *NZ_CP029895.1 | *Salmonella* Typhi strain 311189_252186 plasmid pHCM1 | - | - | - | 7/7 |
| *LC553463.1 | *Escherichia coli* NES6 plasmid pN-ES-6-1 | - | - | - | 7/7 |
| NZ_CP068928.1 | *Klebsiella pneumoniae* strain RIVM_C018860 plasmid pRIVM_C018860_2 | - | - | - | 7/7 |
| MT809700.1 | *Klebsiella pneumoniae* strain 03108465-40B plasmid pKpQIL | - | - | - | 7/7 |

HH=Household
\*Plasmids with antimicrobial resistance genes

**Table 5.** GenBank accession numbers of analyzed *S.* Typhi sequences.

| Isolate No. | BioSample Accession | Whole Genome Sequence Accession | URL |
| --- | --- | --- | --- |
| Ai | SAMN40992305 | JBCHCG000000000 | https://www.ncbi.nlm.nih.gov/nuccore/JBCHCG000000000 |
| Aii | SAMN40992306 | JBCHCF000000000 | https://www.ncbi.nlm.nih.gov/nuccore/JBCHCF000000000 |
| Aiii | SAMN40992307 | JBCHCE000000000 | https://www.ncbi.nlm.nih.gov/nuccore/2725100554 |
| Aiv | SAMN40992308 | JBCHCD000000000 | https://www.ncbi.nlm.nih.gov/nuccore/JBCHCD000000000 |
| Av | SAMN40992309 | JBCHCC000000000 | https://www.ncbi.nlm.nih.gov/nuccore/JBCHCC000000000 |
| Avi | SAMN40992310 | JBCHCB000000000 | https://www.ncbi.nlm.nih.gov/nuccore/JBCHCB000000000 |
| Bi | SAMN40992311 | JBCHCA000000000 | https://www.ncbi.nlm.nih.gov/nuccore/JBCHCA000000000 |
| Bii | SAMN40992312 | JBCHBZ000000000 | https://www.ncbi.nlm.nih.gov/nuccore/JBCHBZ000000000 |
| Biii | SAMN40992313 | JBCHBY000000000 | https://www.ncbi.nlm.nih.gov/nuccore/JBCHBY000000000 |
| Biv | SAMN40992314 | JBCHBX000000000 | https://www.ncbi.nlm.nih.gov/nuccore/JBCHBX000000000 |
| Bv | SAMN40992315 | JBCHBW000000000 | https://www.ncbi.nlm.nih.gov/nuccore/JBCHBW000000000 |
| Ci | SAMN40992316 | JBCHBV000000000 | https://www.ncbi.nlm.nih.gov/nuccore/JBCHBV000000000 |
| Cii | SAMN40992317 | JBCHBU000000000 | https://www.ncbi.nlm.nih.gov/nuccore/JBCHBU000000000 |
| Ciii | SAMN40992318 | JBCHBT000000000 | https://www.ncbi.nlm.nih.gov/nuccore/JBCHBT000000000 |
| Civ | SAMN40992319 | JBCHBS000000000 | https://www.ncbi.nlm.nih.gov/nuccore/JBCHBS000000000 |
| Di | SAMN40992320 | JBCHBR000000000 | https://www.ncbi.nlm.nih.gov/nuccore/JBCHBR000000000 |
| Dii | SAMN40992321 | JBCHBQ000000000 | https://www.ncbi.nlm.nih.gov/nuccore/JBCHBQ000000000 |
| Diii | SAMN40992322 | JBCHBP000000000 | https://www.ncbi.nlm.nih.gov/nuccore/JBCHBP000000000 |
| Div | SAMN40992323 | JBCHBO000000000 | https://www.ncbi.nlm.nih.gov/nuccore/JBCHBO000000000 |
| Dv | SAMN40992324 | JBCHBN000000000 | https://www.ncbi.nlm.nih.gov/nuccore/JBCHBN000000000 |
| Dvi | SAMN40992325 | JBCHBM000000000 | https://www.ncbi.nlm.nih.gov/nuccore/JBCHBM000000000 |
| Dvii | SAMN40992326 | JBCHBL000000000 | https://www.ncbi.nlm.nih.gov/nuccore/JBCHBL000000000 |

Genes conferring multidrug resistance in *S*. Typhi were detected in two different plasmids identified in sub-group 4.3.1.2EA3 (Household D) strains. The antimicrobial resistance genes *sul1*, *dfrA, catA1, aph*(*6*)*-Id, aph(3’’)-Ib and sul2* were detected in a plasmid with sequences corresponding to *S*. Typhi strain 311189_252186 plasmid pHCM1, while *bla_TEM-1_* was detected in a plasmid with sequences corresponding to *E. coli* NES6 plasmid pN-ES-6-1. None of the plasmids in isolates from households A, B and C had identifiable antimicrobial resistance genes.

### Biofilm Formation

We wanted to determine if biofilm-forming ability is correlated with the stage of typhoid fever/carriage, antimicrobial resistance or with the presence of plasmids. Biofilms were examined in gallbladder-simulating conditions. There was varying ability to form biofilms under *in vitro* conditions in the *S.* Typhi strains tested. All isolates formed weak biofilms in absence of both cholesterol and bile (OD_570_ below 0.3) and significantly strong biofilms in presence of cholesterol and 2.5% human bile (Figure 2A and 2B). Differences in biofilm forming ability was observed across the identified genotype 4.3.1 lineages/sub lineages. The sub-lineage 4.3.1.2EA2 formed the strongest biofilms (OD_570_ slightly above 2.0) while sub-lineage 4.3.1.2EA3 formed relatively weak biofilms (OD_570_ below 0.5) even in the presence of cholesterol and human bile (Figure 2A and 2B). The weak biofilm forming isolates, sub-lineage 4.3.1.2EA3, had AMR genes encoded in 2/19 of the plasmids identified in this sub-group. Sub-lineages 4.3.1.2EA2 and 4.3.1.1EA1 had fewer identified plasmids, (14 and 15 respectively) (Table 4), None of these plasmids had identifiable AMR genes, but isolates in these subgroups formed significantly stronger biofilms in the presence of both cholesterol and human bile (Figure 2C). However, there was no statistical significance in biofilm forming ability in strains isolated during the symptomatic vs. asymptomatic stage in each of the three sub-groups of *S.* Typhi (Figure 2D).

**Figure 2.**
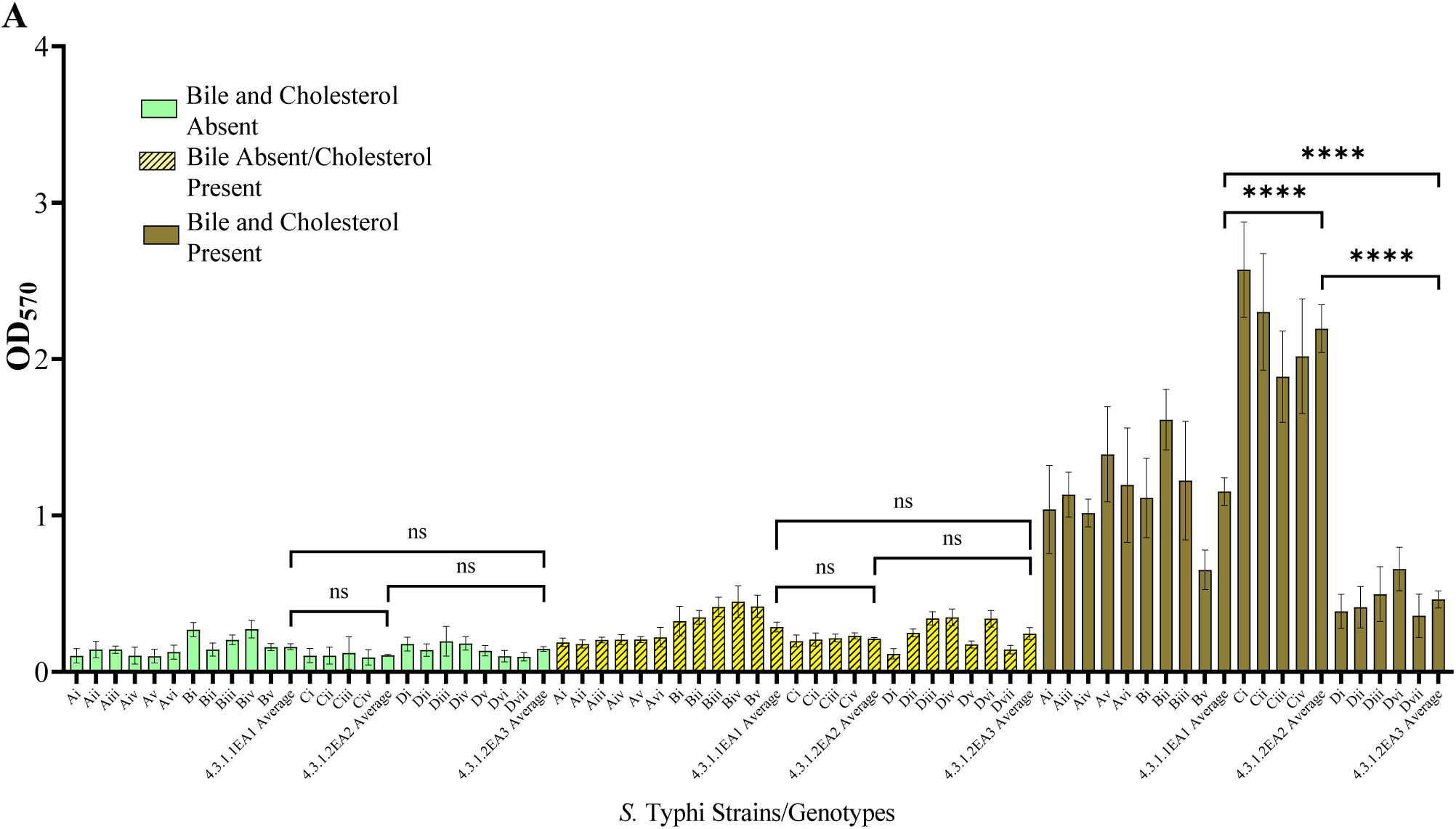
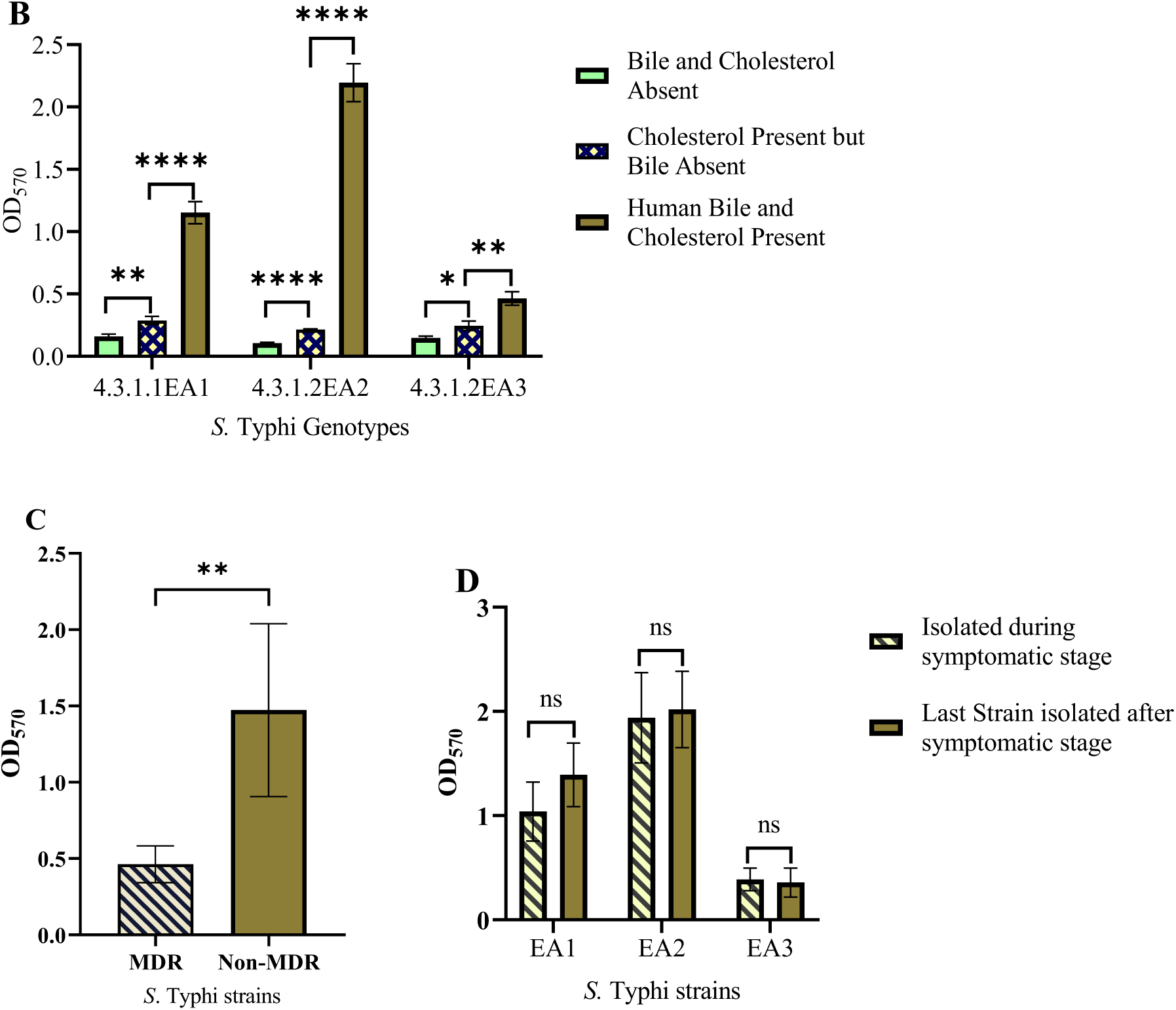
*Salmonella* Typhi biofilms. (A and B) Quantity of biofilms after growth in presence/absence of cholesterol and/or bile. (C) Biofilm formation by MDR *S*. Typhi strains vs. non-MDR strains. (D) Comparison of biofilms formed by *S*. Typhi strains isolated before treatment vs. after treatment. Error bars represent SEM, ****, *P*<0.001; **, *P*<0.05; ^ns^*P*≥0.5.

## Discussion

Although typhoid fever has largely been eliminated in high income countries, it remains a major global public health concern especially among low- and middle-income countries (2). The haplotype 58 (H58), which is associated with antimicrobial resistance has also been reported from other parts of sub-Saharan Africa and Southeast Asia (30,31). In this study, H58 (genotype 4.3.1) was identified as the single genotype shed by four cholelithiasis patients living in a typhoid endemic setting in Nairobi, Kenya. *S*. Typhi H58 is the most dominant genotype in many parts of Southeast and South Asia as well as in East Africa and has spread globally (32,33). Three H58 east African subgroups (4.3.1.1EA1, 4.3.1.2EA2, 4.3.1.2EA3) previously reported circulating in the current study setting by our group (1), were the main lineages/sub-lineages shed by the cholelithiasis patients. The most abundant subgroup was 4.3.1.1EA1 with 11/22 (50%) isolates, originating from individuals living in two different households. In one of these households, an acute case shed an *S.* Typhi belonging to the same sub-group as an asymptomatic household contact who was also diagnosed with gallstones. This suggests possible transmission of the pathogen by the carrier to the household contact (household B). From a different household, a typhoid patient also diagnosed with gallstones continued to shed sub-lineage 4.3.1.2EA2, while in the fourth household, *S.* Typhi sub-lineage 4.3.1.2EA3 strains were isolated from stool samples collected from an acute case after treatment. Unlike the other two sub-groups, the sub-lineage 4.3.1.2EA3 strains had MDR genes, showing resistance to ampicillin, sulfamethoxazole-trimethoprim and chloramphenicol. MDR isolates had more plasmids, 19, compared to the 14 plasmids in non-MDR isolates, from 4.3.1.1EA1 and 15 plasmids from 4.3.1.2EA2. The seven sub-lineage 4.3.1.2EA3 strains had the MDR genes detected in identified plasmids. All 22 *S.* Typhi isolates had point mutations in the QRDR, conferring reduced susceptibility to ciprofloxacin, a drug of choice for treating typhoid fever. There was no variation noted in antimicrobial resistance patterns among strains isolated from the patients in the same household.

Multidrug resistance genes were not detected in 4.3.1.1EA1 and 4.3.1.2EA2 *S*. Typhi genomes, but the strains belonging to these subgroups formed significantly stronger biofilms as compared to the MDR sub-lineage 4.3.1.2EA3 strains. Biofilms act as physical barrier protecting bacteria from killing by antimicrobials including antibiotics. A previous study demonstrated the role of biofilms in protecting *Salmonella* from ciprofloxacin (34). We hypothesize that the 4.3.1.1EA1 and EA2 sub-lineages form better biofilms to counteract the absence of antimicrobial resistance factors, or conversely, that lineage 4.3.1.2EA3 has lost biofilm-related genes because it possesses more plasmids/genes encoding strong antimicrobial resistance. To the best of our knowledge, this is the first study comparing biofilm forming ability in different *S*. Typhi lineages. The mechanism leading to differences in biofilm formation in isolates from the same genotype will need to be further investigated.

Genetic variations were observed in *S*. Typhi from asymptomatic carriers, with a *treB* A383T point mutation being observed in at least one isolate from each of the four households. The *treB* gene codes for PTS trehalose transporter subunit IIBC. As seen in Table 3, household B, some of the mutations observed in the first follow-up isolate were not detected in *S*. Typhi strains isolated during the consecutive follow-ups. However, some mutations were observed in more than one strain isolated from the same patient. From the patient shedding sub-lineage 4.3.1.2EA3 strains, the *tviE* P263S mutation was observed in the fourth isolate and all strains isolated thereafter. This suggests that some of the mutations are maintained in the population during the asymptomatic carriage, while others are not. Although no strain was isolated directly from gallbladder in our study, mutations in the *tviE* gene was also observed in *S.* Typhi gallbladder genome sequences in a previous study (29). The *tviE* gene facilitates the polymerization and translocation of the Vi capsule (35). Vi capsular polysaccharide, an antiphagocytic capsule, covers the surface of *S*. Typhi allowing it to selectively evade phagocytosis by human neutrophils while promoting human macrophage phagocytosis (36). This crucial virulence factor in *S*. Typhi (Vi) also plays a key role in the development of vaccines against typhoid fever (37). However, additional research will be required to understand if this mutation alters the expression of Vi antigen to benefit *S*. Typhi pathogenesis or chronic carriage.

The main limitation in this study is lack of availability of isolates from gallbladder samples from patients shedding *S.* Typhi for comparison with those isolated from stool and blood samples. There were also no *S*. Typhi belonging to other genotypes for comparison purposes.

## Conclusion

The resistance patterns in *S*. Typhi did not change during the duration of asymptomatic carriage in study participants, but these individuals continued to shed and transmit drug resistant strains of this pathogen. Mutations in *S*. Typhi were observed to occur during carriage including those in the Vi antigen locus. Sub-lineages analyzed in this study that were not multidrug resistant, showed the ability to form stronger biofilms than the multidrug resistant strains. This study provides some insights into mutations, drug resistance and biofilm formation during typhoid carriage, and this information may be used to influence public health approaches aimed at reducing carriage and transmission of *S*. Typhi.

## Methods

### Source of Bacteria strains

The whole genome sequences of 22 *S*. Typhi strains isolated from blood and stool samples of six patients living in four different households in Mukuru informal settlement, a typhoid endemic area in Nairobi, Kenya, were analyzed. Nineteen of these strains were from four patients who were also diagnosed with cholelithiasis, of whom, three had typhoid symptoms and continued to shed *S.* Typhi after treatment. The presence of gallstones was confirmed through an ultrasound scan, the primary imaging modality performed by a radiologist used to evaluate patients suspected of having gallbladder disease. One of the patients with gallstones was asymptomatic but shedding *S*. Typhi and living in the same household (household B) with an acute typhoid fever case (not diagnosed with gallstones). In a different household (household A), a study participant (household contact) not diagnosed with cholelithiasis shed *S*. Typhi once. The index case from this household was diagnosed with cholelithiasis and continued to shed *S*. Typhi after treatment with antibiotics. Laboratory methods on isolation and identification of the isolates are as described in our previous publication (38).

### Whole Genome Sequencing

DNA extracted from *S*. Typhi strains using GenElute™ Bacterial Genomic DNA Kit (Missouri, United States) was prepared for whole genome sequencing by SeqCoast Genomics using an Illumina DNA Prep tagmentation kit and unique dual indexes. Sequencing was performed on the Illumina NextSeq2000 platform using a 300-cycle flow cell kit to produce 2×150bp paired reads as previously described (39). PhiX control, 1-2%, was spiked into the run to support optimal base calling. Read demultiplexing, read trimming, and run analytics were performed using DRAGEN v3.10.12, an on-board analysis software on the NextSeq2000. Raw sequencing data generated during this study are available in the National Center for Biotechnology Information (NCBI) public data, BioProject ID PRJNA1101423.

### Genome Assembly and Annotation

Quality-trimming of the reads was done using Trimmomatic (40). Read error was corrected using SPAdes (41). Short (and long, if available) reads were assembled into contigs using SPAdes (wrapped in Unicycler). Read mapping was done using Bowtie2 and SAMtools (wrapped in Unicycler) (42–44) (https://github.com/rrwick/Porechop). Pilon (wrapped in Unicycler) was used in polishing of each assembly (45). Gene prediction and functional annotation was performed using BAKTA (46). The annotation pipeline was as follows; prediction of protein-coding genes using Prodigal, tRNA identification using tRNAscan-SE (47), tRNA and tmRNA identification using Aragorn (48), prediction of rRNA sequences using Infernal and the Rfam database (49), CRISPR prediction using PILER-CR (50), antimicrobial resistance gene identification using AMRFinderPlus (51), prediction of signal peptides using DeepSig (52), prediction of transposases using ISFinder (53) and computation of codon usage biases for each amino acid.

### Genotype Identification and Bacteria Clustering

Identification and clustering of *S*. Typhi genotypes was performed using Pathogenwatch (https://pathogen.watch/), a web-based platform with several different components (54,55). The platform provides compatibility with *S*. Typhi typing information for MLST (56), in silico serotyping (SISTR) (57), and a SNP genotyping scheme (GenoTyphi) (55). *S.* Typhi Assemblies (in fasta format) were uploaded to the platform https://pathogen.watch/upload/fasta for the analysis.

### Antimicrobial Susceptibility Testing

Antimicrobial susceptibility testing was performed using the disk diffusion technique for all antimicrobials commonly used in Kenya for typhoid fever treatment including ampicillin (10 µg), tetracycline (30 µg), co-trimoxazole (25 µg), chloramphenicol (30 µg), amoxicillin-clavulanate (20/10 μg), cefpodoxime 30 µg, ceftazidime (30 µg), ceftriaxone (30 µg), cefotaxime (30 µg), azithromycin (15 μg), ciprofloxacin (5 µg), nalidixic acid (10 µg), kanamycin (30 μg) and gentamicin (10 μg). The diameter of the zone of inhibition was measured after 18-24 hours and results were interpreted according the Clinical and Laboratory Standards Institute (CLSI) guidelines for *Salmonella* (CLSI 2023).

### Screening of Antimicrobial Resistance Genes

NCBI Antimicrobial Resistance Gene Finder Plus (AMRFinderPlus) (https://github.com/ncbi/amr/wiki), was used to identify acquired antimicrobial resistance genes and known resistance-associated point mutations in *S*. Typhi assembled nucleotide sequences (51). The results were compared with phenotypic susceptibility data.

### Variant Calling

To identify genetic variations between the longitudinal clinical isolates, *S*. Typhi strains isolated before each typhoid index case was treated with antibiotics were used as the reference genome, and were compared with those isolated after treatment (follow-up isolates), and/or those isolated from household contacts using breseq (58). The variant calling pipeline was as follows; quality-filtering of raw reads using Trimmomatic (40), mapping of reads against a reference genome (first strain isolated from the index case), analysis of possible mutations based on mapping data, identification of mutations and graphical and tabular summaries of mutation profile across samples (42).

### Plasmid Identification

To identify plasmids in the isolated *S.* Typhi strains, a plasmid detection tool PLASMe was used (https://github.com/HubertTang/PLASMe). The tool uses the alignment component in PLASMe to identify closely related plasmids while diverged plasmids are predicted using order-specific Transformer models (59).

### In *Vitro* Biofilm Formation Assays

Because of the importance of biofilm formation on gallstones in chronic carriage (17,19) the biofilm forming ability of all the 22 human *S*. Typhi isolates were tested under gallbladder simulating conditions. *S*. Typhi biofilms were grown on non-treated polystyrene 96-well plates (Corning, Kennebunkport, ME). To simulate growth conditions on gallstones, wells in two plates were pre-coated with cholesterol by adding a solution of 5 mg/mL in 1:1 isopropanol:ethanol and air-dried overnight. A pure colony of *S*. Typhi on an XLD agar plate was cultured in Tryptone Soy Broth (TSB). Overnight (O/N) cultures in broth were normalized to OD_600_=0.8, diluted 1:2500 in TSB or TSB containing 2.5% human bile, and 100 µL/well were dispensed into the plates. The plates were incubated at 25°C in a Fisherbrand™ nutating mixer (Thermo Fisher Scientific; Hampton, NH) at 24 rpm for 96 hours. Media (TSB or TSB containing bile) was changed after every 24 hours for consistent *S*. Typhi biofilm growth. Plates were emptied on the fourth day and washed twice before heat fixing at 60°C for 1 hour. The biofilms were stained using a crystal violet solution and acetic acid (33%) used to elute crystal violet before reading the OD_570_. GraphPad prism 9.5 was used to analyze the biofilm formation results. One way Analysis of variance (ANOVA) was used to test level of significance in biofilm formation between the different *S*. Typhi sub-lineages and in different conditions, i.e., biofilms in absence of cholesterol and bile, in cholesterol coated plates in absence of bile, and in presence of cholesterol and bile. Student’s t-test was used to test level of significance in biofilm formation in strains isolated before treatment vs last strains shed by the patient, P-values less than 0.05 (P<0.05) were considered significant.

## Data Availability

The whole genome sequences from all the 22 *S*. Typhi isolates were deposited to the National Center for Biotechnology Information (NCBI), GenBank accession numbers are shown in the table below (Table 5).

## Acknowledgements

This research was financially supported by the National Institute of Health (NIH) National Institute of Allergy and infectious Diseases (Grants: R01 AI099525 and R01 AI116917).

